# Prior infection by seasonal coronaviruses does not prevent SARS-CoV-2 infection and associated Multisystem Inflammatory Syndrome in children

**DOI:** 10.1101/2020.06.29.20142596

**Authors:** Isabelle Sermet-Gaudelus, Sarah Temmam, Christèle Huon, Sylvie Behillil, Vincent Gajdos, Thomas Bigot, Thibaut Lurier, Delphine Chrétien, Marija Backovic, Agnès Moisan-Delaunay, Flora Donati, Mélanie Albert, Elsa Foucaud, Bettina Mesplées, Grégoire Benoist, Albert Faye, Marc Duval-Arnould, Célia Cretolle, Marina Charbit, Mélodie Aubart, Johanne Auriau, Mathie Lorrot, Dulanjalee Kariyawasam, Laura Fertitta, Gilles Orliaguet, Bénédicte Pigneur, Brigitte Bader-Meunier, Coralie Briand, Vincent Enouf, Julie Toubiana, Tiffany Guilleminot, Sylvie van der Werf, Marianne Leruez-Ville, Marc Eloit

## Abstract

**Background:** Children have a lower rate of COVID-19, potentially related to cross-protective immunity conferred by seasonal coronaviruses (HCoVs). We tested if prior infections with seasonal coronaviruses impacted SARS-CoV-2 infections and related Multisystem Inflammatory Syndrome (MIS).

**Methods:** This cross-sectional observational study in Paris hospitals enrolled 739 pauci or asymptomatic children (HOS group) plus 36 children with suspected MIS (MIS group). Prevalence, antigen specificity and neutralizing capability of SARS-CoV-2 antibodies were tested. Antibody frequency and titres against Nucleocapsid (N) and Spike (S) of the four seasonal coronaviruses (NL63, HKU1, 229E, OC43) were measured in a subset of seropositive patients (54 SARS-CoV-2 (HOS-P subgroup) and 15 MIS (MIS-P subgroup)), and in 118 matched SARS-CoV-2 seronegative patients (CTL subgroup).

**Findings:** SARS-CoV-2 mean prevalence rate in HOSP children was 11.7% from April 1 to June 1. Neutralizing antibodies were found in 55·6% of seropositive children, and their relative frequency increased with time (up to 100 % by mid-May). A majority of MIS children (25/36) were SARS-CoV-2 seropositive, of which all tested (n=15) had neutralizing antibodies. On average, seropositive MIS children had higher N and S1 SARS-CoV-2 titres as compared to HOS children. Patients from HOS-P, MIS-P, and CTL subgroups had a similar prevalence of antibodies against the four seasonal HCoVs (66·9 −100%). The level of anti-SARS-CoV-2 antibodies was not significantly different in children who had prior seasonal coronavirus infection.

**Interpretation:** Prior infection with HCoVs does not prevent SARS-CoV-2 infection and related MIS in children. Children develop neutralizing antibodies after SARS-CoV-2 infection.

**Evidence before this study:** Children seem to be less likely affected by SARS-CoV-2 infection and clinical course of COVID-19 is less severe than in adults. As those asymptomatic or mildly symptomatic children are underdiagnosed and their viral loads are comparable to those of adults, they may act as an asymptomatic reservoir for the spread of the virus. One explanation of the difference between the adult and the pediatric infectious profile might be that infection with seasonal human coronaviruses, which is very frequent from a very young age, could lead to cross protective immunity. We searched in PubMed, MedRxiv and BioRxiv for publications from inception to June 15, 2020, using the terms “COVID-19, SARS-CoV-2, children, serology, Kawasaki, Corona Virus”.

**Added value of this study:** SARS-CoV-2 mean prevalence rate was 11.7% from April 1 to June 1 and neutralizing antibodies were found in 55% of the tested seropositive children. Among patients with a Multisystem Inflammatory Syndrome, Kawasaki-like disease, 70% were SARS-CoV-2 seropositive and had neutralizing antibodies. COVID-19 and MIS attack rates, and anti-SARS-CoV-2 antibodies titres were not significantly impacted by prior seasonal coronavirus infection.

**Implications of all the available evidence:** Prior infection by seasonal coronaviruses does not prevent SARS-CoV-2 infection and associated Multisystem Inflammatory Syndrome in children As antibodies against seasonal coronaviruses are very frequent and as these viruses circulate efficiently in human populations every winter, our results question to what extent the concept of herd immunity based on circulating antibodies can be applied to seasonal coronaviruses and possibly SARS-CoV-2.

## Introduction

COVID-19 is due to SARS-CoV-2, a betacoronavirus subgenus Sarbecovirus^1^, which has expanded worldwide since its emergence in China at the end of 2019. Observations indicate that children are less likely to develop the disease and that the clinical course of COVID-19 in children is less severe than in adults^2–4^. Accordingly, children represent only 0.6-2.3% of confirmed cases in China and 0.8-5.2% outside China outside context of household ^2,5,6^. As asymptomatic or mildly symptomatic children are underdiagnosed, and their viral loads are comparable to those of adults, children may act as an asymptomatic reservoir for the spread of the virus to their adult and elderly relatives^7,8^, albeit with low efficacy^9^. Children’s susceptibility to infection might also be low^5^ and the paediatric cohort may represent a pool of “immune naïve” population. Differences in susceptibility profiles for children and adults might be driven by infections with seasonal human coronaviruses (HCoVs), which are very frequent at a very young age^10^, and could lead to cross-protective immunity in children. This may be mediated either by cross-binding or cross-neutralizing antibodies^11^, or by T cell responses that target epitopes shared by SARS-CoV-2 and HCoVs^12,13^. Indeed, it was recently shown that CD4+ T cells of unexposed subjects (sampled before the pandemic) recognized SARS-CoV-2.

Despite a low frequency of respiratory symptoms, cases of Multisystem Inflammatory Syndrome (MIS) have been reported in children that were infected by SARS-CoV-2 or were in contact with COVID-19 patients^14,15^. MIS shares similarities with classic Kawasaki disease but displays different prominent clinical signs including cardiogenic shock or myocarditis^15^. As for other post infectious diseases^16^, it is possible that a low antibody response to SARS-CoV-2, or cross-reactive antibodies without any neutralizing capability, facilitate immune-dependent enhancement following re-exposure, potentiated by a specific genetic background^17,18^. Interestingly, a domain of the SARS-CoV-2 spike protein which binds with high affinity to T cells may act as a super antigen, and trigger excessive adaptive immune responses^19^.

The aim of this study was to analyse the impact of endemic seasonal coronaviruses on SARS-CoV-2 infection in children. This was performed in a large cohort of children aged 0 – 18 years, hospitalized in Paris. To measure if prior infections with HCoVs (detected by antibody responses against two major antigens, S and N) conferred protection towards SARS-CoV-2 infection, we analysed their frequency in SARS-CoV-2 positive children as compared to SARS-CoV-2 negative matched controls. We also analysed SARS-CoV-2 and seasonal HCoVs humoral responses of patients with MIS regarding antibody targets and functional neutralizing activity. Our study is the first to analyse in depth the typology of humoral responses to SARS-CoV-2 in children, and provides evidence that prior infections by seasonal coronaviruses has no significant impact on SARS-CoV-2 infection or related MIS disease in children.

## Methods

### Study setting and data collection

Paediatric patients aged 0 – 18 years consulting or hospitalized for at most 4 days in a paediatric tertiary health care department of the Assistance Publique-Hôpitaux de Paris between April 1, 2020, and June 1, 2020 were included in this prospective multicentric observational seroprevalence study. Children hospitalized for COVID-19 were excluded. Sera were also collected from March 1 to March 31 from patients consulting for regular follow-up or hospitalized in emergency in one of the hospitals (Necker Enfants Malades Hospital). The definition of a suspected case for COVID-19 was based on recommendations from the European Center for Disease Prevention and Control (ECDC)^20^. We also included in this study during the same period patients presenting with a MIS disease, as defined by the American Heart Association^21^.

We recorded the history of suspected COVID-19 cases based on a standardized study-specific form. We collected data on clinical symptoms consistent with COVID-19 occurring from December 2019 until up to 7 days before enrolment. Demographic information, relevant epidemiological history (e.g. international travel or contact with an infected person or a suspected case), comorbidities, reasons for hospitalization were collected.

### Ethics

The local Ethics (CERAPHP Paris V) approved this study (IRB registration: #00011928). Patients and/or their parents/guardians were informed about the study

### Serological tests

For SARS-CoV-2 prevalence, we used a LIPS (Luciferase Immunoprecipitation System) test as previously reported^22^, which identifies antibodies (Ab) targeted to the S1 and S2 domains of the Spike (S) and to the Nucleoprotein (N). For the neutralization assay we used a viral pseudotype-based assay and a neutralization test using live SARS-CoV-2. In a fraction of samples, we assessed by LIPS tests, antibodies to the nucleoprotein and to the full spike ectodomain in a pre-fusion conformation for SARS-CoV-2, and the four human coronaviruses (HKU1, NL63, OC43, and 229E). Detailed technical information together with sensitivity and specificity evaluations are given respectively in **Supplementary Material S1 and S2**.

### Statistical analysis

We did not detect any difference in terms of age, sex ratio, and main comorbidities between patients recruited in March (n=133; single centre study) and afterwards (n=642; multicentric seroprevalence study) (**Supplemental Table 1**). We therefore present the demographic and clinical data of the whole cohort of children enrolled. Cases were grouped based on the results of serology testing and clinical presentation. Data were assessed for normal distribution using the Kolmogorov-Smirnov test. Continuous variables were presented as mean (SD) and compared using the Student t test. Categorical variables were compared using Chi2 or Fisher’s exact tests, as appropriate. Statistical analyses were conducted with Excel or GraphPad Prism 8 (GraphPad Software, LLC). Principal Component Analysis was performed to identify the serological profile according to SARS-CoV-2 Abs and seasonal HCoV Abs. Data were processed with R 3.6.3 using GGPlot2 with GGally for matrices of plots, and ggfortify for PCA plots packages. Two-sided p value of <0.05 was considered significant.

## Results

### 1-Prevalence and clinical presentation of SARS-CoV-2 infection

In total, 775 children, mean (SD) age 8 (5·6) years were enrolled in the study (**Figure 1**). One hundred six (13·6%) were below 15 years of age. The reason for consultation or hospitalization was regular follow-up for 58%, surgery for 11·7% and medical emergency for 15% (HOS population). Among those, 17 patients (2·1%) were hospitalized for neurological disorders including encephalitis (n=4), cerebellitis (n=5), polyradiculoneuritis (n=7), and labyrinthitis (n=1). Thirty-six other patients (4·6%) presented with a MIS and among those, 25 were hospitalized in Intensive Care Unit because of signs of shock, and 28 developed myocarditis.

**Figure 1:**
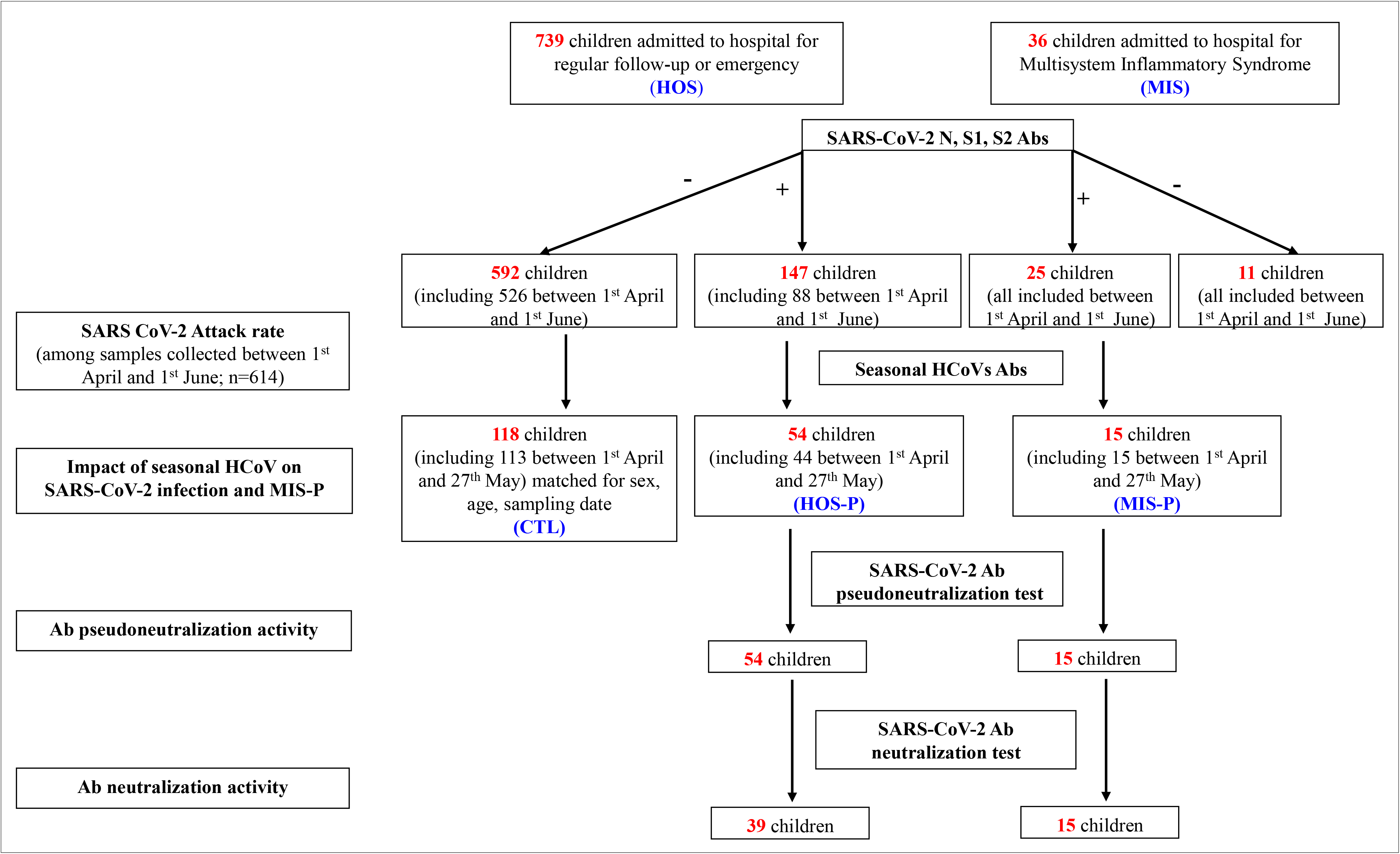
Flowchart of inclusion of children (n=775)

The apparent prevalence of seropositive children was in the range 10% − 15% between 6^th^ April and 1^st^ June, except on the week starting 11^th^ May where it reached 27%. The patients with MIS were mainly detected from the last week of April to mid-May (**Figure 2**). The seropositive patients with neurological disorders were hospitalized over the whole study period.

**Figure 2:**
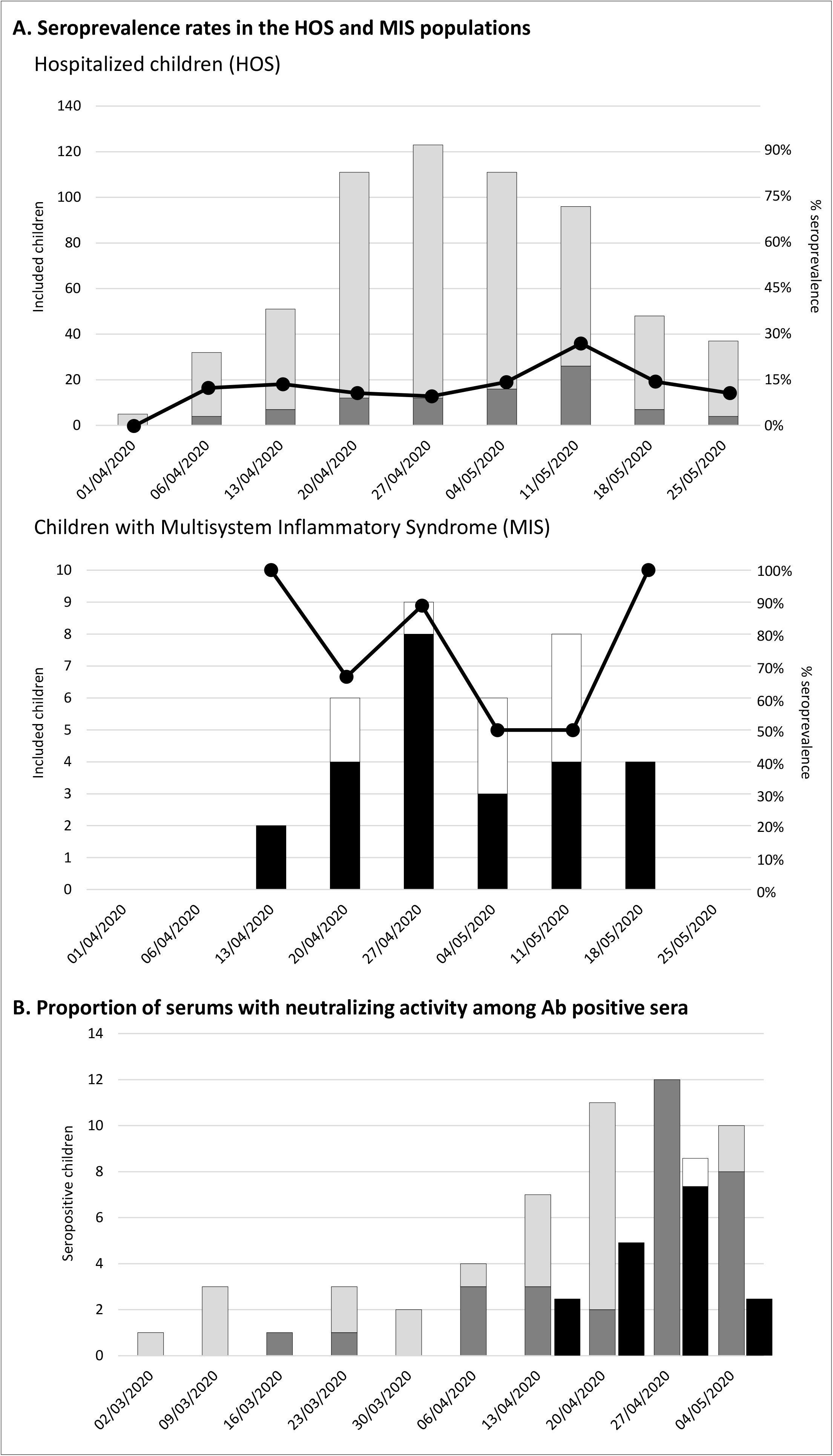
Evolution of the prevalence of antibodies and neutralizing activity to SARS-CoV-2 in children during the epidemic. A) Seroprevalence rates in the HOS and MIS populations. Seronegative (light grey) and seropositive (dark grey) children according to the week of sampling are presented. The black curve describe the percentage of seroprevalence over the time. B) Proportion of serums with neutralizing (PNT) activity among Ab positive sera. The number of PNT positive (respectively in dark grey and black for HOS-P and MIS-P patients) and negative (in light grey and white for HOS-P and MIS-P patients, respectively) sera is presented over the time.

**Table 1** presents the demographic and clinical characteristics of 594 SARS-CoV-2 seronegative patients and 172 seropositive patients enrolled between 1^st^ March to 1^st^ June. The seropositive children consisted of patients hospitalized for MIS (MIS-P; n=25) or for any other reason (HOS-P; n=147). The comparison between seronegative and seropositive patients among the whole cohort of HOS and MIS populations did not show any significant differences for age, sex ratio, reasons for hospitalization and main comorbidities, apart for SARS-CoV-2 seropositive MIS patients, who did not report underlying chronic diseases and were all hospitalized in emergency units. There was a significantly higher frequency of household contact with suspected SARS-CoV-2 infection (without testing) among the seropositive patients. History of contact with a suspected COVID-19 household significantly increased the risk for positive serology, even in case of asymptomatic infection (OR 2.25, 95% CI [1.3; 3.9]). Among the 25 contacts of seropositive patients, 23 were parents.

**Table 1:**
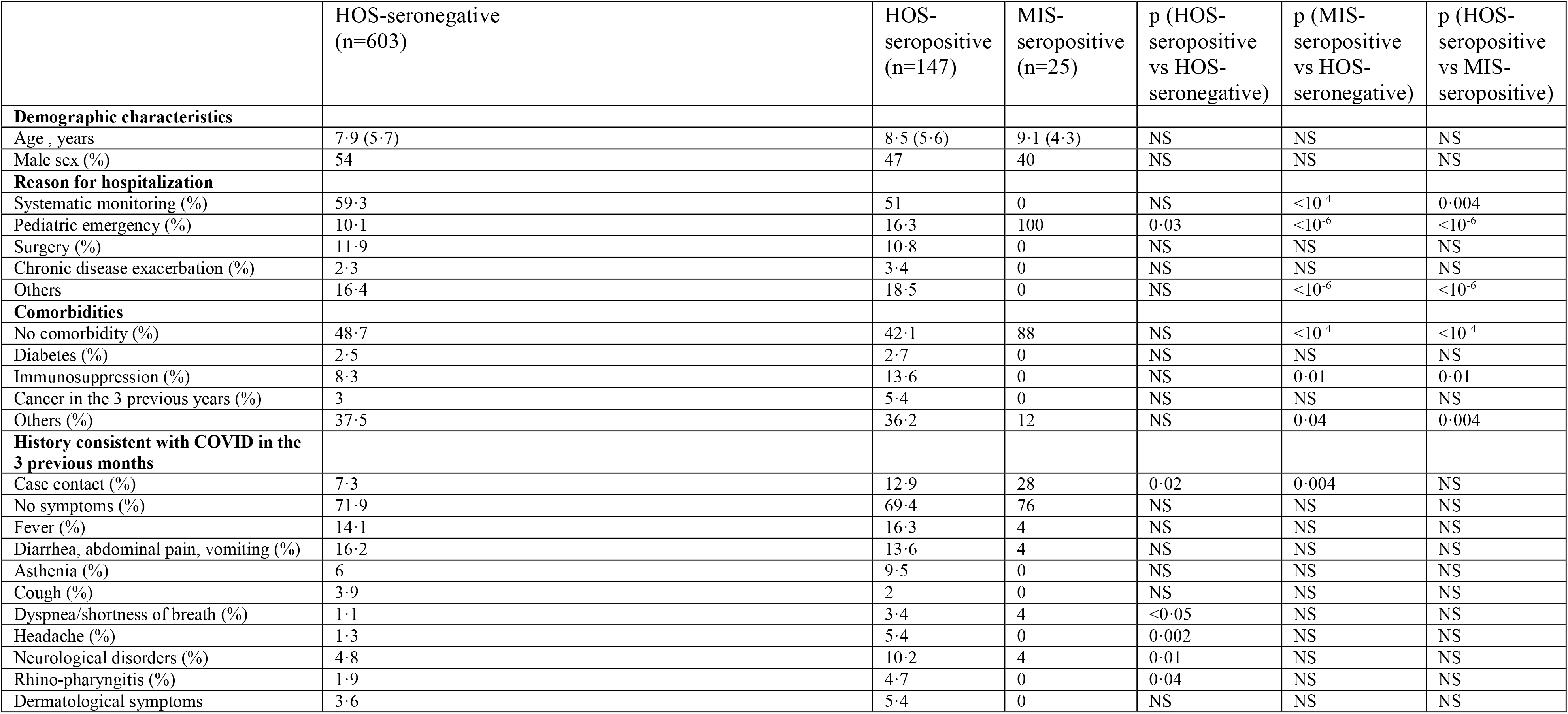
Demographic and clinical characteristics of children with SARS-CoV-2 Ab versus negative controls. HOS stands for patients who did not develop a Multi-systemic Inflammatory Syndrome (MIS). Data as mean (SD) or %.

The seropositive children (HOS-P and MIS-P) did not show an age-dependent specific distribution (**Supplemental Figure S3)**. More than 70% of them (121 out of the 172) did not report any history consistent with COVID-19 during the preceding weeks. The only symptoms that were marginally but significantly reported in the previous months in seropositive children were headache, shortness of breath and rhino-pharyngitis. Neurological disorders were significantly more frequent in the seropositive group (13 children out of 17).

### 2-Profiling SARS-CoV-2 antibody responses in HOS-P and MIS-P patients

HOS-P patients Ab profile was characterized by a dominant S2 response compared to responses to S1, N and to the full S ectodomain (**Figure 3**). The HOS-P children with neurological symptoms showed N, S1 and S2 responses similar to those of the HOS-P group without neurological symptoms (**Supplemental Figure S4**) and were analysed together.

**Figure 3:**
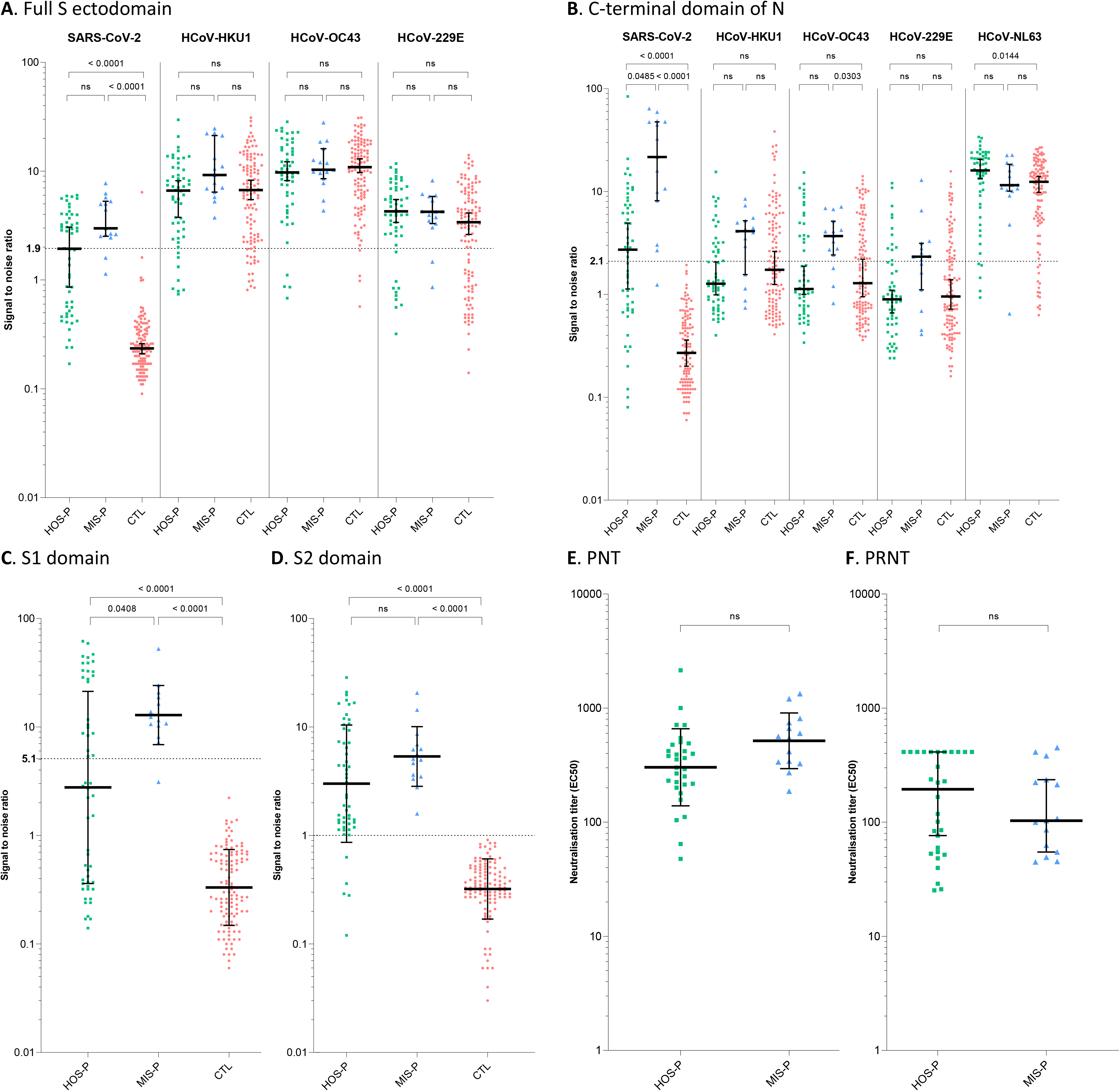
SARS-CoV-2 and seasonal HCoV Ab responses in HOS-P, MIS-P and CTL children. Ab directed against S (panel A), N (panel B), S1 (panel C) and S2 (panel D) are described for each group of patients (HOS-P in green, MIS-P in blue, and CTL in pink). Level of neutralizing activity assessed by pseudo-neutralization (panel E) or plaque reduction neutralization (panel F) is presented for HOS-P and MIS-P groups.

MIS-P patients showed a distinct Ab profile directed against N, S1 and S2 altogether (**Figure 4**). Levels of N and S1 Abs to SARS-CoV-2 were significantly higher in the MIS-P than in the HOS-P group (**Figure** **3C-D**).

**Figure 4:**
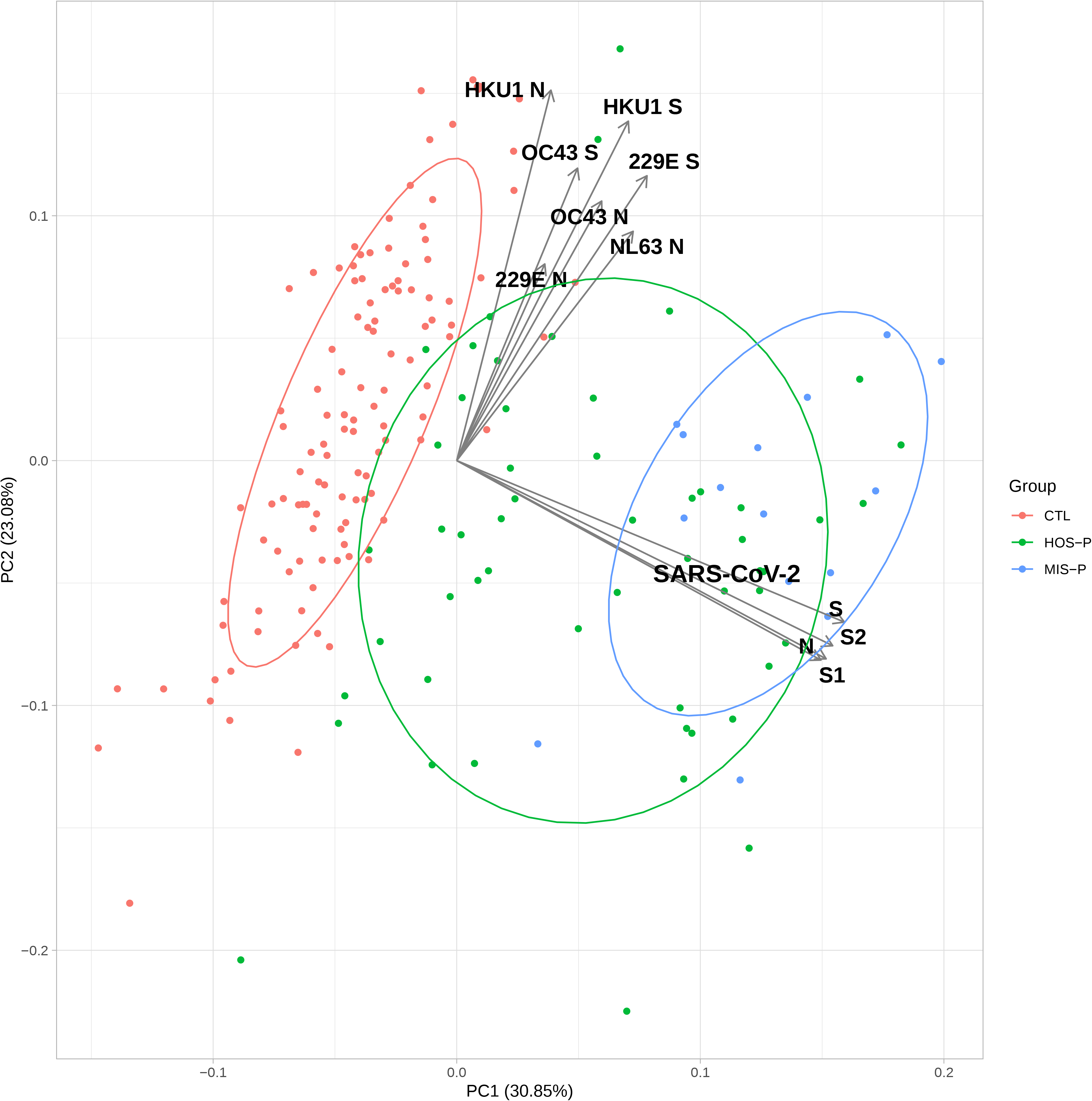
Principal Component Analysis of antibodies against SARS-CoV-2 S, S1, S2, N and seasonal HCoV S and N antigens. The PCA shows (A) the distribution of all Ab against SARS-CoV-2 mainly correlated with the x-axis and those corresponding to seasonal HCoVs with the y-axis; and (B) Abs against seasonal HCoVs do not separate HOS-P and MIS-P from CTL children. This demonstrates the independence of Ab responses between SARS-CoV-2 and HCoVs.

### 3-Neutralization activity of the Ab

To investigate the neutralization activity, we first performed a pseudo-neutralization assay in the subset of 54 HOS-P and 15 MIS-P patients (**Figure 1**). Overall, 55·6% of HOS-P and 100% of MIS-P children showed a neutralizing activity. Interestingly, the fraction of HOS-P children whose Abs displayed a pseudo-neutralizing activity (PNT+) increased with time (**Figure 2B**), from 18% to 38% during March and April to 100% at the beginning of May. For a fraction of these children, we investigated the correspondence between PNT and live SARS-CoV-2 neutralizing activity determined in a plaque reduction test (PRNT) (**Table S6**). All tested sera positive in PNT (n=28) were PRNT+ and 2 out of the 11 PNT-negative sera were also PRNT+. In total, 73·7% of HOS-P tested sera showed a neutralizing activity. Neutralizing titres of PNT+ and PRNT+ sera of MIS-P patients were similar to those of HOS-P patients (**Figure 3**).

### 4-Relationship between SARS-CoV-2 and seasonal HCoV infections

We compared the prevalence of anti-N and -S antibodies against the four seasonal HCoVs in a subpopulation of children among the HOS-P (n=54), MIS-P (n=15) and CTL (n=118) groups (**Figure 1**). Prevalence rates for the two betacoronaviruses (HKU1 and OC43) and the two alphacoronaviruses (229E and NL63) were similar for all viruses between the CTL and HOS-P groups. They were also similar for HKU-1, 229E and NL63 in the MIS-P group, whereas Abs to OC43 N were more frequent in the MIS-P group (73%) than in the CTL group (39.4%), which was not paralleled by the anti-S response that was near 100% in the two groups (**Table 2**).

**Table 2:**
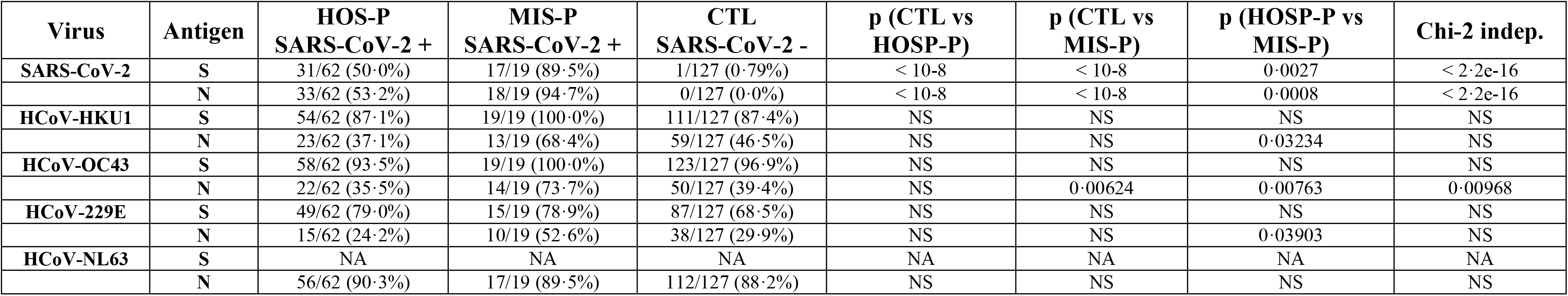
prevalence of antibodies to seasonal HKU1, OC43, NL63, 229E and SARS-CoV-2 spike and nucleoprotein in HOS-P, MIS-P and CTL children. NA: not applicable. NS: non-significant.

We reasoned that prior infection with SARS-CoV-2, identified by antibody responses, could induce cross-reactive immune effectors, like CD4+ and CD8+ T cells and neutralizing antibodies. This would diminish or impair SARS-CoV-2 antibody responses, which should therefore be lower in patients with high anti-HCoV Ab levels. Alternatively, previous HCoV infections could prime the immune system resulting, upon SARS-CoV-2 infection, in a boost of the antibody response against shared epitopes or in facilitation of infection. Such boost should lead to higher anti-SARS-CoV-2 Ab levels in HCoV positive patients. There was no significant difference between HOS-P and CTL patients regarding antibody levels to the four seasonal HCoVs (**Figure 3**). This demonstrates that humoral responses between seasonal coronaviruses and SARS-CoV-2 were not related. This is confirmed by the PCA analysis which showed that the patient groups (HOS-P, MIS-P and CTL) were only clustered by SARS-COV-2 Abs and not by seasonal HCoV Abs. (**Figure 4**). This suggests that the risk of SARS-CoV-2 infection and of related MIS disease were not influenced by prior seasonal HCoV infection. In support to this observation, there was an association, for each virus, between all S and N antibody titres within each of the three groups of patients (CTL, HOS-P and MIS-P). This correlation was stronger for SARS-CoV-2 than for seasonal HCoVs. (**Supplemental Figures S5 and S6**) Importantly, there was no significant association between SARS-CoV-2 and seasonal HCoV quantitative Ab responses (**Supplemental Figures S5 and S6**). MIS-P and HOS-P groups did not differ for Abs titers against seasonal HCoVs, except for OC43-N Abs that were increased in MIS-P children, but this tendency was not confirmed by the OC43-S Ab response **(Figure 3A-B)**. This was coherent with the Abs prevalence study **(Table 2)**..

## Discussion

This cross-sectional prospective multicentric study provides to the best of our knowledge the largest report on COVID-19 in the paediatric population outside of China. We enrolled nearly 800 children during the first three months of the COVID-19 epidemic in Paris. From April to May, the prevalence rate of Abs against SARS-CoV-2 was in the range of 10-15%. We noticed an outlier peak of incidence during the week of 11 May, which was likely associated with an increase of vigilance of physicians following the information of severe clinical presentations in children. This prevalence rate is in contrast with previous epidemiological cohort studies, based on RT-PCR, where children represent less than 2% of diagnosed cases^24-23^. These numbers are compatible with the prevalence estimated in Paris^24^ despite the specificity of the population of children recruited in university hospitals. Here, in a large cohort of children less than 15 years old, we demonstrate that (i) a substantial proportion of children can become infected and (ii) the risk is not related to age. In nearly 15% of the cases, a contact with a parent suspected of COVID-19 was identified, which increased by 2·5 fold the risk to be infected. Very importantly, more than 50% of the seropositive children did not report any symptoms, a proportion similar to that recently reported in adults^25^. The reminders reported mild and non-specific symptoms such as headache, rhino-pharyngitis and shortness of breath. This confirms previously published data showing that COVID-19 is less severe in children than in adults^26^. The largest study in children published so far that described SARS-CoV-2 infection in 2,143 Chinese children also reported asymptomatic infection or mild symptoms such as fever, cough, a sore throat, sneezing, myalgia and fatigue^27^. Altogether, these results and ours underline that most children remain undiagnosed because of asymptomatic infections, which makes them potential drivers of virus spread^28^.

This is to our knowledge the first study profiling the humoral immune response in children experiencing pauci-symptomatic infection by SARS-CoV-2. We showed that around half of SARS-CoV-2 positive sera present a significant neutralizing activity based on two independent assays (based on lentivirus pseudo-typed with the SARS-CoV-2 spike or based on live virus). Interestingly, this rate increased up to 100% at the end of the observation period on mid-May, almost 2 months after the peak of the epidemic. As reported by others^29^, this suggests that appearance of neutralizing antibodies is delayed relative to initial seroconversion. This is in contrast to severe COVID-19 forms^22,30^, where neutralizing antibody responses against the immunodominant S viral protein are elicited as soon as after two weeks of infection at higher frequency and titres. Importantly, considering these data, it must be underlined that quantitative correlates of protection are currently unknown, which makes it difficult to relate the neutralizing titres to a clinically relevant effect.

To investigate reasons explaining decreased severity of SARS-CoV-2 infection in children, we studied the impact of prior infections with seasonal HCoVs on the risk of infection by SARS-CoV-2. Antibodie were considered as an evidence of past infection by HCoVs, the intensity of the antibody response reflecting partly the degree of replication within the host. However, this represents a limitation as cases of infection have been reported to be seronegative as revealed by isolated T-cell responses^31^. Seasonal HCoVs include alphacoronaviruses (229E and NL63) and betacoronaviruses lineage A (OC43 and HKU1), which primarily replicate in the respiratory tract and mostly cause common colds^32^. These viruses show a worldwide distribution and multiple HCoV infections in various combinations are common^10,32–34,35^. Infection takes place in very early childhood and seroprevalence studies show very high prevalence rates^36^, up to 100% in adult populations^10,37,38^. It is therefore very likely that infection by HCoVs preceded infection by SARS-CoV-2 in our cohort. Previous seasonal HCoV infections could have an impact on SARS-CoV-2 replication. Indeed, antibodies are unlikely to act as primary effectors of protection, as there is no or very low^11^ cross neutralization between these coronaviruses, but antibodies serve as an indicator of underlying cellular responses. We found no evidence of cross-protective immunity linked to previous infection by seasonal HCoVs. First, similar seasonal HcoV prevalence was found in SARS-CoV-2 positive versus negative patients. Second, on a quantitative side, there was no significant correlation between SARS-CoV-2 and any HCoV antibody titres, whatever the antigen considered (S or N). On the contrary, the level of SARS-CoV-2 antibodies to N and S were correlated, which corresponds to a good internal control. This was also the case for the N and S responses for each HCoV, but to a lesser extent. We hypothesized that multiple infections by seasonal HCoVs would boost Ab responses against shared epitopes that are more frequent in the nucleoprotein than in the spike, and that this would lead to a decrease of correlation between N and S responses over time. When analysing the S and N responses to HCoVs, we found no obvious difference in N to S correlations in CTL patients as compared to the HOS-P group, suggesting that infection with SARS-CoV-2 did not significantly boost pre-existing antibody responses to HCoVs N and S.

This lack of HCoV/SARS-CoV-2 cross-protection contrasts with the recent demonstration of pre-existing immune effectors recognizing SARS-CoV-2 in subjects sampled before the SARS-CoV-2 pandemic. Moreover, a very sensitive cytometric assay reported frequent low levels of cross-reacting anti-S IgGs, mainly targeting the SARS-CoV-2 S2 domain of the spike^11^. Although those Abs neutralized entry of SARS-CoV-2 S-pseudo-typed lentiviruses in HEK-293T cells mediated by the spike protein, the clinical relevance of the SARS-CoV-2 pseudo-neutralisation test is questionable, because the mechanism of entry did not involve the ACE2 receptor of the virus. Moreover, T-helper cells detected in healthy subjects also recognized the C-terminal part of S (that contains the S2 subunit) but not the receptor-binding domain (RBD) which belongs to S1^13^. In contrast, our results clearly show that cross-reactive antibodies directed against endemic seasonal HCoVs and underlying cross-reacting CD4+ T-cells do not seem to confer any significant protection against SARS-CoV-2 infection. This could be explained by low identity between coronaviruses of important targets such as the RBD. Importantly, this also suggests that potentially cross-reactive CD8+ T cells, which should be elicited upon seasonal HCoV infections as is the case following SARS-CoV-2 infection^39^, are not able to significantly contribute to protection against SARS-CoV-2 infection.

We analysed the SARS-CoV-2 Ab profile in 25 MIS cases associated with a positive SARS-CoV-2 Ab response. We found higher S1 and N responses, but not an increased neutralizing capacity as compared to HOS-P patients who experienced an asymptomatic or pauci-symptomatic infection (**Figure 2A**). This was not the case for the beta-(OC43) or alpha-(229E and NL63) coronaviruses Ab responses, suggesting that the increased SARS-CoV-2 response is not a non-specific feature triggered by inflammation. Furthermore, the lack of cross-reactivity between anti-S1 Abs of the different viruses^40^ does not favour the hypothesis of SARS-CoV-2 infection boosting pre-existing HCoV immunity in MIS patients. As a whole, our data do not support that previous HCoV infection facilitates SARS-CoV-2 infection and MIS-related disease.

Neutralizing tests for the four seasonal HCoVs were not available but some extrapolation can be done based on SARS-CoV-2 neutralization and LIPS results, because equivalent LIPS tests were used for all viruses. As the SARS-CoV-2 results show that up to 100% of LIPS positive sera sampled one to two months post-infection had neutralizing activity, it can be anticipated that most HCoV positive sera would also neutralize corresponding HCoVs. This means that, in a perspective of herd immunity, most of the general population would have antibodies neutralizing seasonal HCoVs, among other immune effectors. As common colds due to seasonal HCoVs are experienced repeatedly, this leads to questioning whether coronavirus immune responses induce a long-term clinically protective response. Our results therefore pose a doubt regarding the humoral protective response against SARS-CoV-2 in a perspective of herd immunity, even when the prevalence of antibodies will be high in the population.

The strengths of the study are the high number of well documented paediatric cases including MIS cases and the extensive exploration of Ab responses to SARS-CoV-2, their neutralizing activity, and the correlation with seasonal HCoV immune responses. We acknowledge that infection rates are probably biased in this cohort of hospitalized children, despite the fact that we focused as much as possible on regular follow-up or COVID-19 unrelated emergencies. In this cohort, we observed no correlation between the Ab responses against SARS-CoV-2 and seasonal HCoVs. Although cellular and local immune defenses were not directly tested, our results do not provide evidence for cross-protective immunity or facilitation linked to previous infections by seasonal HCoVs on the risk of developing a SARS-CoV-2 infection or a MIS disease once infected.

In conclusion, our results show that 11.7 % of children admitted in Parisian hospitals have Abs against SARS-CoV-2 and that these Abs are able to neutralize SARS-CoV-2 *in vitro*. As seasonal HCoVs circulate efficiently in the human population despite very high antibody prevalence, our results point to the limits of herd immunity applied to seasonal coronaviruses and maybe SARS-CoV-2.

## Data Availability

The data of the study will be available on request

## Author contribution

Conceptualization and Methodology:

Cohort management and sample collection:

Serological and seroneutralisation assays:

Data assembly and manuscript writing:

Funding acquisition:

Supervision:

All authors reviewed and approved the final version of the manuscript.

## Acknowledgments

We thank Thierry Rose and Yves Janin for providing us the luciferase reagents, Evelyne Dufour and Stephane Petres for their help in the production of recombinant proteins, Pierre Charneau and François Anna for testing the sera with the pseudoneutralization test and the ICAReB platform for giving us access to a collection of sera collected before the epidemic. We thank Olivier Schwartz for providing us with the raw data of the S-Flow technique for the sera analyzed in Grzelak et al., 2020. We thank Quentin Leclech for help with data analysis, and Zakary M’Sakny, Mathis Crespin, Felix Wolfram, Victor Zetlaoui (enthusiastic MD Students), Deborah Rechard, Dr Michaela Semeraro and Dr Emeline Roy who helped for data collection. We thank Simon Cauchemez for critical lecture of the manuscript.

